# Lymphocytopaenia is associated with severe SARS-CoV-2 disease: A Systematic Review and Meta-Analysis of Clinical Data

**DOI:** 10.1101/2020.04.14.20064659

**Authors:** Robin Brown, Jane Barnard, Eva Harris-Skillman, Bronwen Harbinson, Beata Dunne, Jonathan Drake, Sophie Roche, Edward Harris, James Gunnell, Joshua Frost, Brian Angus, Susanne H Hodgson

**Author notes:** **Corresponding Author Details:** Mr Robin Brown, Harris Manchester College, Mansfield Rd, Oxford, OX1 3TD.

## Abstract

**Background:** Most patients infected by SARS-CoV-2 have favourable outcomes, however some develop severe disease which may progress to acute respiratory distress syndrome, multi-organ failure, and death. Markers that could predict patients at risk of poor outcomes would be extremely useful clinically. Evidence has emerged that low lymphocyte count is associated with increased disease severity.

**Methods:** We performed a systematic review and meta-analysis to assess the association between lymphocyte count and severity of SARS-CoV-2 associated clinical disease.

**Results:** Seven papers were included in the meta-analysis. These papers included data from 2083 patients, 25% (n=521) with severe SAR-CoV-2 disease and 75% (n=1562) with non-severe SAR-CoV-2 disease. Heterogenicity was seen in the definition of severe disease. Metanalysis produced metamedians of 1×10^9^/L (95% CI 1-1.1) and 0.7×10^9^/L (95% CI 0.63-0.8) lymphocytes for patients with non-severe and severe disease respectively (*p-*value of p=0.006 *Wilcoxon test*). Calculation of metamedians from the two papers classifying severe disease according to death alone gave 1.1 1×10^9^/L lymphocytes (95% CI 1.0-1.1) for ‘survivors’ (n=163) and 0.63 1×10^9^/L lymphocytes (95% CI 0.60-0.63) for ‘non-survivors’ (n=253) of SAR-CoV-2 disease.

**Conclusions:** Lower lymphocyte counts are significantly associated with more severe disease in patients with SARS-CoV-2 infection. Lymphocytopenia may therefore be useful laboratory measure to allow prognostication of patients presenting with SARS-CoV-2 infection.

## INTRODUCTION

Since it was first described in Wuhan, Hubei Province, China, in November 2019^1^, the novel human coronavirus Severe acute respiratory syndrome coronavirus 2 (SARS-CoV-2) has spread globally. At the time of writing, there are 1,051,635 confirmed cases, with a total of 56,985 deaths attributed to the virus worldwide^2^. SARS-CoV-2 causes coronavirus disease 2019 (COVID-19)^2^, a viral pneumonia characterised by a high fever, persistent dry cough, dyspnoea and fatigue^3–5^. Clinical manifestations range from asymptomatic disease to severe respiratory distress^6^. There are as yet no clearly efficacious drug treatments for COVID-19, and management is largely supportive^6^.

Although most patients with COVID-19 have a favourable prognosis, some develop dyspnoea, hypoxaemia, and rapidly progress to life-threatening acute-respiratory distress syndrome (ARDS) and end-organ failure^4^. Given the propensity of this disease to cause rapid decline, early recognition of patients who are likely to develop severe disease would enable more effective treatment planning and resource allocation. A clinically validated prognostic scoring system would therefore be extremely useful clinically.

While the clinical symptoms and signs of COVID-19 have been extensively reviewed in published literature^1,3,4,14-17^, clinical and laboratory characteristics that could inform prognostication have been less well studied. One laboratory marker of interest is lymphocyte count, with lymphocytopaenia associated with poor disease outcomes^7,8^. Lymphocytopaenia is already recognised as an independent marker of mortality in community-acquired and ‘ICU-acquired’ pneumonias, and is associated with increased disease severity^9,10^. Lymphocyte count can be assessed with a simple blood sample, making it a pragmatic, widely available measure for clinical prognostication.

This work seeks to further define the degree of correlation between lymphocytopaenia and severity of COVID-19 disease through a systematic review and meta-analysis of the available literature.

## METHODS

### Literature Search

This protocol follows the recommendations established by the Preferred Reporting Items for Systematic Reviews and Meta-Analyses (PRISMA) statement^11^.

We conducted a systematic review using Embase and Medline, searching for articles relating to the current COVID-19 outbreak from November 2019 until 30 March 2020. The following search terms were used: [[COVID-19] OR [2019-nCoV] OR [SARS-CoV-2]] AND clinical characteristics. The reference lists of identified articles were checked to identify any relevant missed papers.

Results of the initial search were screened by title and abstract and then inclusion and exclusion criteria applied to the full texts of relevant articles (Table 1). Included articles compared severe and non-severe cases of COVID-19, where severe disease was classified as one or more of: RR>30, SaO2<93%, paO2/FiO2<300mmHg, ARDS, ICU admission, mechanical ventilation, or death. Only papers containing adult patients with confirmed COVID-19 based on positive RT-PCR for SARS-CoV-2 and/or positive CT findings were included. Publications were required to report a lymphocyte count with measurements presented as a median with inter-quartile range (IQR). Articles were excluded if; appropriate information was not reported, patients had a pre-existing condition or were taking medication that would affect lymphocyte count, or if there was no English language version of the paper available.

**Table 1:** Inclusion and Exclusion Criteria

|  | Inclusion Criteria | Exclusion Criteria |
| --- | --- | --- |
| Date range | Post-COVID-19 outbreak: Nov 2019 – present |  |
| Language |  | No English language version of paper available |
| Study design | Comparison of SEVERE and NON-SEVERE cases of COVID19 where severe is classified as one/more of:<br>• RR>30, SaO2<93%, paO2/FiO2<300mmHg<br>• ARDS<br>• ITU admission<br>• Mechanical ventilation<br>• Death<br>'Non-Severe' disease is cases not reaching these criteria |  |
| Patient characteristics | Adults $\geq 18$ years old<br>Confirmed COVID-19 based on one or both of:<br>• positive RT-PCR for SARS-CoV-2<br>• Positive CT findings | Any other infection, rheumatological, or inflammatory conditions, or any other condition/medication that would cause immunosuppression or itself change lymphocyte count/CRP |
| Biological parameters | • A reported lymphocyte count<br>• Values presented as median (IQR)<br>• Values reported separately (not compositely) for SEVERE vs NON-SEVERE groups. | Self-reported studies, qualitative data |

Nine authors (J.B, R.B, J.D, J.F, J.G, B.H, E.H, E.HS, S.R.) screened and evaluated the identified articles. Each paper was evaluated by two independent authors and any discrepancies were resolved by a third author (Figure 1). The following features were extracted for pooled estimation: number of patients in the study, number of severe cases, number of non-severe cases, lymphocyte count median and IQR.

**Figure 1:**
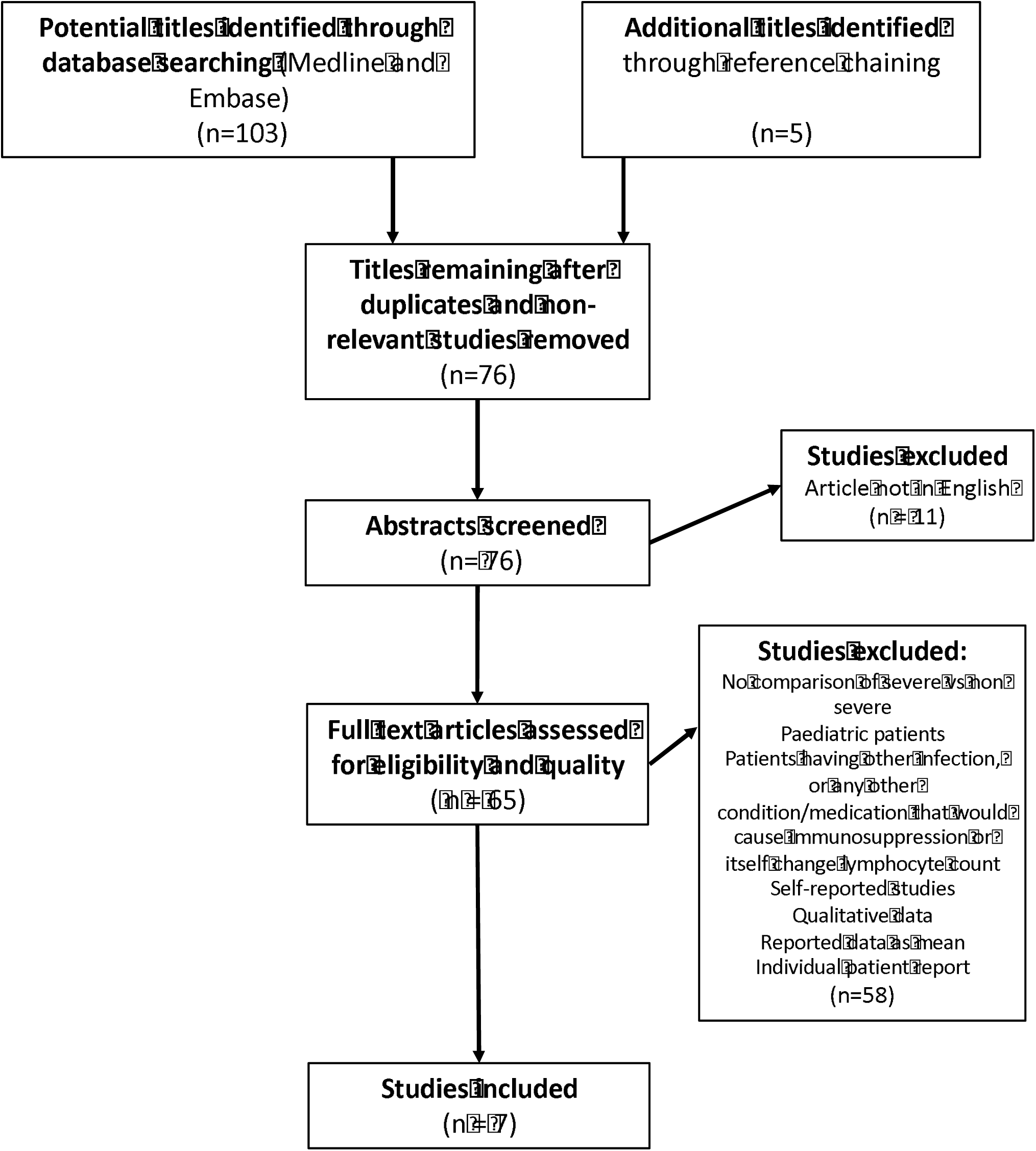
Summary of Paper Review Process.

### Statistical Analysis

The data were presented using the median and interquartile range, a non-normal distribution was assumed, and non-parametric tests were performed. Independent two-group Wilcoxon tests were used to compare median lymphocyte counts between severe and non-severe populations, and to calculate confidence intervals. The R package ‘metamedian’^12^ was used to produce a meta-median lymphocyte count for each case type. Confidence intervals were produced to the 95% confidence level and *p-*values of *p*<0.05 were taken to be significant.

## RESULTS

After review, seven papers were included in the meta-analysis^3,13–18^ (Figure 1 & Table 2). These papers included data from 2083 patients, 25% (n=521) with severe SAR-CoV-2 disease and 75% (n=1562) with non-severe SAR-CoV-2 disease. Data extracted from these papers are summarised in Figure 3. The majority of papers defined severe disease according to clinical criteria related to hypoxia and need for intensive care^3,13–16^, however two papers defined severe disease only as infection resulting in death (Table 2)^1718^. Metanalysis of lymphocyte data from all seven papers gave metamedians of 1×10^9^/L (95% CI 1-1.1) and 0.7×10^9^/L (95% CI 0.63-0.8) lymphocytes for patients with non-severe and severe disease respectively (*p=0*.*006 Wilcoxon test*) (Table 3).

**Table 2:**
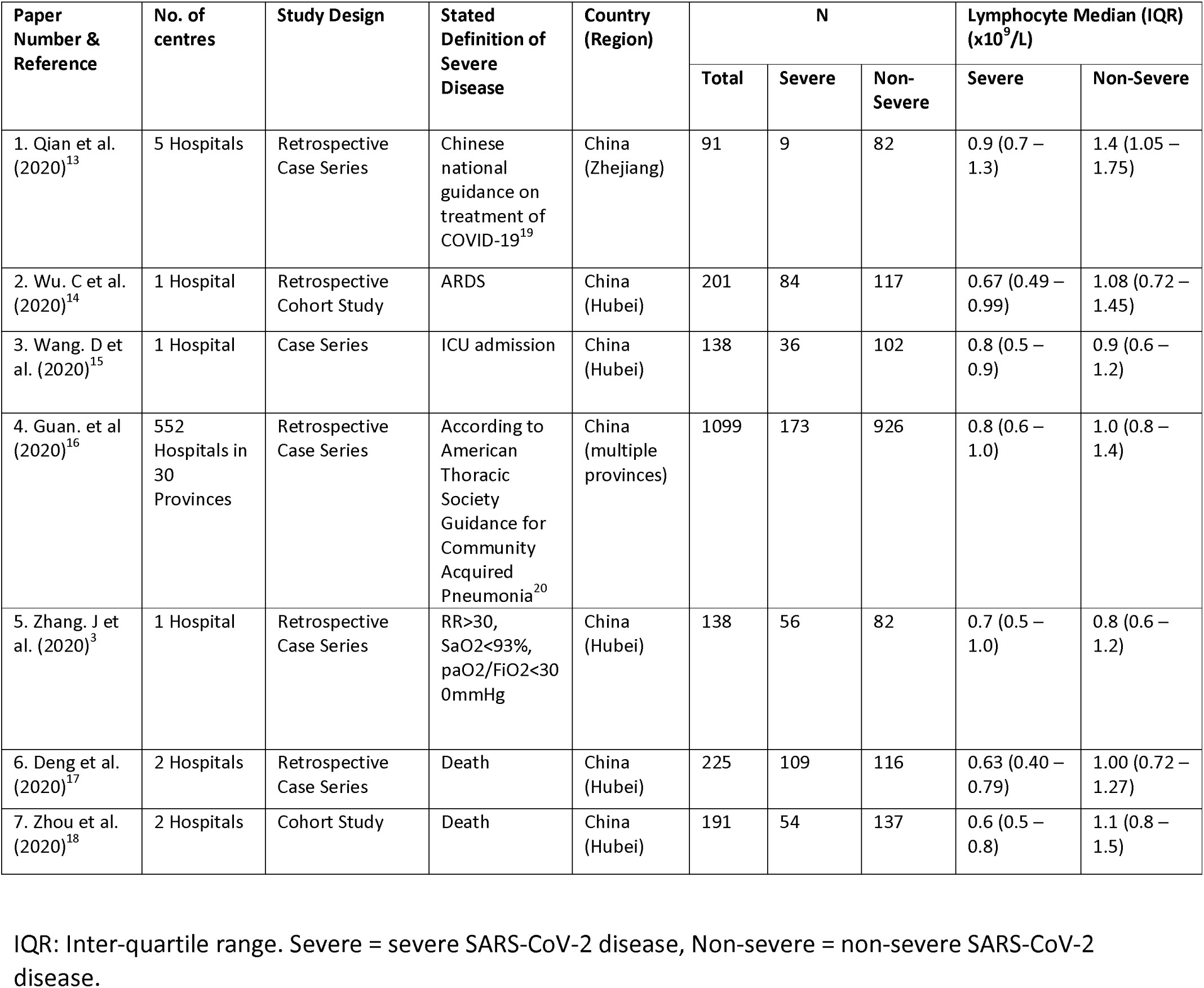
Characteristics of Papers Included in the Meta-analysis of Association between Severity of SARS-CoV-2 infection and Lymphocyte Count

**Table 3:** Lymphocyte Metamedians calculated for patients with severe and non-severe SARS-CoV-2 disease

|  | Lymphocyte Metamedian (95% CI) (x10<sup>9</sup>/L) |  | P-value |
| --- | --- | --- | --- |
|  | Severe Disease | Non-Severe Disease |  |
| **All Papers (n=7)<sup>3, 12-17</sup>** | 0.7 (0.63 – 0.8)<br>*n=521* | 1 (1 – 1.1)<br>*n=1562* | *0.006* |
| **Excluding papers using death as sole criterion for severe disease (n = 5)** | 0.8 (0.67 – 0.8)<br>*n=358* | 1 (0.8 – 1.4)<br>*n=1309* | *0.044* |
| **Papers using death as sole criterion for severe disease (n=2)** | 0.63 (0.6-0.63)<br>*n=163* | 1.1 (1-1.1)<br>*n=253* | * |

Metamedian values were calculated using the R package ‘metamedian’<sup>18</sup> using Wilcoxon test. \* *p value* not calculated. n = number of patients.

Given the heterogenicity in definition of severity of disease, we repeated the metanalysis excluding data from two papers that defined severe disease as mortality, as some survivors in these papers may have the met criteria for severe disease used in other publications. This analysis included data from 1,667 patients; 358 with severe disease and 1309 with non-severe disease. Metanalysis produced metamedians of 1×10^9^/L (95% CI 0.8-1.4) and 0.8×10^9^/L (95% CI 0.67-0.8) lymphocytes for non-severe and severe patient groups respectively (Table 3) (*p=0*.*044 Wilcoxon test*).

Calculation of metamedians of lymphocyte count from the two papers classifying severe disease according to death alone^1718^ gave 1.1 1×10^9^/L lymphocytes (95% CI 1.0-1.1) for ‘survivors’ (n=163) and 0.63 1×10^9^/L lymphocytes (0.60-0.63) for ‘non-survivors’ (n=253) of SAR-CoV-2 disease (Table 3).

## DISCUSSION

Our meta-analysis assessing association between lymphocyte count and severity of SARS-CoV-2 disease has revealed that lower lymphocyte counts are significantly associated with worse disease and poorer prognosis. Whilst the definition of severe disease varied between studies, a clear association was demonstrated, with lowest lymphocyte counts seen in the most severe SARS-CoV-2 disease that resulting in death. To our knowledge, this is the first time this correlation has been formally reported.

There are several limitations to be considered in this finding. Firstly, we do not know the temporal relationship between a decline in lymphocyte count and the onset of more severe disease, and so it is unclear how early this measure could be used to predict disease severity. It is also of note that as the largest multicentre study did not list the centres used in their analysis, so there is a possibility that some patients were counted twice in papers included in our metanalysis^15^. However, despite these limitations, this review confirms what anecdotally clinicians have widely observed, that lymphocytopenia is associated with severe COVID-19 disease^6^.

Lymphocytopaenia was demonstrated to be a feature of SARS (SARS-CoV) infection^19^. Autopsy studies revealed this was due to targeted toxicity to lymphocytes and lymphoid tissue^21^. Given the genetic similarity to SARS-CoV, it may be that similar pathology is a feature of infection with SARS-CoV-2^22^. The correlation between lymphocyte count and severity of COVID-19 infection complements two recent meta-analyses by Lippi *et al*. 2020 which showed a correlation between thrombocytopenia and procalcitonin levels and severity of COVID-19 disease^23,24^. Lymphocyte count could therefore be used alongside these and other laboratory measures to assess the severity of illness in patients with SARs-CoV-2 infection and potentially to predict outcome.

## Data Availability

All the data used in the study is available in the publications used for the meta-analysis

## ACKNOWLEDGEMENTS

SHH is an Academic Clinical Lecturer funded by the National Institute of Health Research and a Research Fellow at St Peter’s College, University of Oxford.

## ETHICS

There are no ethical considerations for this paper

## AUTHOR CONTRIBUTIONS

Robin Brown – Study design, Literature search, Data Collection, Paper writing, corresponding author.

Jane Barnard – Data Collection, review of manuscript

Eva Harris-Skillman – Data Collection, Paper writing, review of manuscript

Bronwen Harbinson – Data Collection, Paper writing, Table preparation, review of manuscript

Beata Dunne – Statistical Analysis, review of manuscript

Jonathon Drake – Data Collection, Table preparation, review of manuscript

Sophie Roche – Data Collection, Paper writing, Figure preparation, review of manuscript

Edward Harris – Data Collection, review of manuscript

James Gunnell – Data Collection, Table preparation, review of manuscript

Joshua Frost – Study design, Data collection, review of manuscript

Brian Angus – review of manuscript

Susanne H Hodgson – Paper writing, review of manuscript

## EXCLUSIVE LICENCE

“I, Robin Andrew Campbell Brown, The Corresponding Author of this article contained within the original manuscript which includes any diagrams & photographs within and any related or standalone film submitted (the Contribution”) has the right to grant on behalf of all authors and does grant on behalf of all authors, a licence to the BMJ Publishing Group Ltd and its licencees, to permit this Contribution (if accepted) to be published in the BMJ and any other BMJ Group products and to exploit all subsidiary rights, as set out in our licence set out at: http://www.bmj.com/about-bmj/resources-authors/forms-policies-and-checklists/copyright-open-access-and-permission-reuse.”

I am one author signing on behalf of all co-owners of the Contribution.

## PPI

Patients or the public WERE NOT involved in the design, or conduct, or reporting, or dissemination plans of our research

## TRANSPARENCY DECLARATION

The lead author, Robin Brown, affirms that this manuscript is an honest, accurate, and transparent account of the study being reported; that no important aspects of the study have been omitted; and that any discrepancies form the study as planned (and, if relevant, registered) have been explained.

## DISSEMINATION DECLARATION

Not applicable for this study.

